# Severe infection among young infants in Dhaka, Bangladesh: effect of case definition on incidence estimates

**DOI:** 10.1101/2024.09.24.24314067

**Authors:** Alastair Fung, Cole Heasley, Lisa G. Pell, Diego G. Bassani, Prakesh S. Shah, Shaun K. Morris, Davidson H. Hamer, Mohammad Shahidul Islam, Abdullah Al Mahmud, Eleanor Pullenayegum, Samir K. Saha, Rashidul Haque, Md. Iqbal Hossain, Chun-Yuan Chen, Abby Emdin, Karen M. O’Callaghan, Miranda G. Loutet, Shamima Sultana, S. M. Masum Billah, S. M. Abdul Gaffar, Enamul Karim, Sharika Sayed, Sultana Yeasmin, Md. Mahbubul Hoque, Tahmeed Ahmed, Shafiqul A. Sarker, Daniel E. Roth

**Author notes:** **Corresponding author:** Alastair Fung, Division of Paediatric Medicine, The Hospital for Sick Children, 555 University Ave, Toronto, Ontario, M5G 1X8, Canada.

## Abstract

**Introduction:** Heterogeneity in definitions of severe infection, sepsis and serious bacterial infection (SBI) in young infants limits the comparability of randomized controlled trials (RCTs) of infection prevention interventions. To inform the design of severe infection prevention RCTs for young infants in low-resource settings, we estimated the incidence of severe infection in an observational cohort of Bangladeshi infants aged 0-60 days and examined the effect of variations in case definitions on incidence estimates.

**Methods:** In 2020-2022, 1939 infants born generally healthy were enrolled at two hospitals in Dhaka, Bangladesh. Severe infection cases were identified through up to 12 scheduled community health worker home visits from 0-60 days of age or through caregiver self-referral. The primary severe infection case definition combined physician documentation of standardized clinical signs and/or diagnosis of sepsis/SBI, plus either a positive blood culture or parenteral antibiotic treatment for ≥5 days. Incidence rates were estimated for the primary severe infection definition, the World Health Organization (WHO) definition of possible SBI, blood culture-confirmed infection, and five alternative severe infection definitions.

**Results:** Severe infection incidence per 1000 infant-days was 1.2 (95% CI 0.97-1.4) using the primary definition, 0.84 (0.69-1.0) using the WHO definition of possible SBI, and 0.026 (0.0085-0.081) using blood culture-confirmed infection. One-third of cases met criteria for the primary severe infection definition through physician diagnosis of sepsis/SBI rather than the standardized clinical signs, and 85% of cases were identified following caregiver self-referral despite frequent scheduled study visits.

**Conclusions:** Severe infection incidence in young infants varied considerably by case definition. A severe infection definition that requires physician documentation of standardized clinical signs may miss a substantial proportion of cases identified by physician diagnosis of sepsis/SBI. In settings where health facilities are accessible, frequently scheduled home assessments by study personnel to identify severe infection in infants may not be necessary.

**What is already known on this topic:**

- Researchers aiming to design a randomized controlled trial (RCT) for severe infection prevention or treatment in young infants require a clinically precise and feasible case definition of severe infection.
- A previous systematic review of neonatal sepsis definitions used in RCTs identified a diverse range, including culture-confirmed sepsis, a combination of clinical signs and culture-confirmation, and a combination of clinical signs and laboratory investigation results.
- Incidence estimates of various severe infection case definitions that can be operationalized in low- and middle-income countries (LMICs) are needed to determine the feasibility of using these definitions in severe infection prevention and treatment RCTs for young infants in these settings.

**What this study adds:**

- We provide incidence estimates of severe infection in young infants born generally healthy in Dhaka, Bangladesh, during the first 60 days of age using case definitions based on different combinations of clinical signs, antibiotic treatment and microbiologic criteria.
- We demonstrate that the incidence estimates of severe infection in young infants vary considerably depending on whether a permissive or stringent case definition is adopted.
- We also demonstrate that in this study, most severe infection cases were identified following caregiver self-referral rather than during scheduled home assessments by study personnel.

**How this study might affect research, practice or policy:**

- Our findings may inform the design of future severe infection prevention RCTs in young infants in LMICs by 1) providing incidence estimates of various candidate case definitions, and 2) supporting the planning of optimal outcome surveillance systems that balance the identification of severe infection cases with operational costs.

## INTRODUCTION

Severe infections, including sepsis, contribute to a large burden of disease early in life and result in significant morbidity and mortality in young infants (<2 months).^1–3^ The burden of sepsis in young infants is particularly high in low- and middle-income countries (LMICs).^4,5^ Randomized controlled trials (RCTs) of preventive and therapeutic interventions are essential for developing evidence-based guidelines for young infant management, but conducting such trials requires a case definition of severe infection relevant to this age group and the trial setting. Various definitions have been used to denote severe infection in young infants, including sepsis and serious bacterial infection (SBI). A major challenge limiting the comparability of RCTs of severe infection prevention and treatment interventions for young infants is the marked heterogeneity in definitions used in studies and guidelines.^6,7^ A positive sterile site culture is often regarded as the gold-standard definition of bacterial sepsis in infants but these are often not available due to limited microbiological resources (blood culture bottles, lack of laboratory capacity, etc.), inadequate sample volume, antibiotic administration prior to sample collection, or low yield despite suitable sample collection without prior antibiotic administration.^8^ A systematic review of neonatal sepsis definitions used in RCTs identified a diverse range; the most common was culture-confirmed sepsis (27% of definitions), followed by clinical signs and culture-confirmation (23%), and then clinical signs and laboratory investigation results (20%).^9^

In LMICs, sick young infants are often initially assessed in the community or at first-level health facilities where access to physicians and laboratory investigations is limited. In a cohort of young infants brought to a hospital or outpatient clinic for an acute illness, the Young Infants Clinical Signs Study Group developed a clinical sign-based algorithm (seven signs) to identify severe illnesses requiring urgent hospital management.^10^ The World Health Organization (WHO) has since adapted these seven signs to define clinical infection syndromes in young infants, including possible serious bacterial infection (pSBI), that identify sick infants at risk of SBI or a severe illness requiring referral and/or empiric antibiotic treatment.^11,12^ The criteria have evolved over time, but currently, pSBI denotes one or more of the following: fever, hypothermia, poor feeding, history of convulsions, lethargy, elevated respiratory rate or severe chest indrawing.^11^

When designing RCTs for young infant severe infection prevention in LMICs, the use of a relatively permissive case definition (e.g., pSBI) will result in higher event rates and, therefore, require a lower minimum sample size, compared to the use of more stringent case definitions (e.g., culture-confirmed sepsis). However, the use of relatively permissive definitions may dilute intervention effects, meaning that experimental interventions that truly prevent bacterial infections may be misleadingly observed to have similar overall event rates as the control group. Conversely, when using a highly stringent severe infection case definition, the outcome may be so rare that the trial may be too costly or considered unfeasible to complete within a reasonable timeframe.

The Synbiotics for the Early Prevention of Severe Infections in Infants (SEPSiS) observational cohort study (NCT04012190) has several aims including the investigation of the incidence of severe infection in a cohort of young infants (0-60 days of age) born generally healthy in Dhaka, Bangladesh. The findings from this cohort were intended to be used to design severe infection prevention trials in generally healthy young infants. The primary case definition of severe infection selected for the SEPSiS study aimed to balance permissiveness and stringency while ensuring that the definition could be feasibly operationalized in the study setting. The case definition of severe infection combined physician documentation of standardized clinical signs and/or diagnosis of sepsis/SBI, plus either a positive sterile site culture or hospitalization with an intention to treat with parenteral antibiotics for ≥5 consecutive days. The term *infection* was used rather than *sepsis* to acknowledge that many infants will not manifest overt signs of sepsis (i.e., signs of systemic inflammatory response syndrome), and the qualifier *severe* implies that the infection may be life-threatening or cause significant morbidity if untreated, such that a young infant would conventionally be admitted to the hospital and empirically treated with parenteral antibiotics.

We aimed to estimate the incidence of severe infection and non-injury-related death up to 60 days of age in Bangladeshi infants born generally healthy, and to examine the effect of variations in severe infection case definitions on incidence estimates. We also examined the proportion of young infants with severe infection identified through various referral pathways as part of the study design. The results may inform the design of future severe infection prevention trials in young infants in LMICs and provide insight into optimal surveillance systems that balance the identification of cases with operational costs.

## METHODS

### Study design and participants

Between November 25, 2020, and February 18, 2022, potential mother-infant pairs were screened for eligibility at two government healthcare facilities in Dhaka city: Maternal and Child Health Training Institute (MCHTI) in Azimpur and Mohammadpur Fertility Services and Training Centre (MFSTC). Additional details of study sites are provided in **Supplemental File, Section S1**. Generally healthy infants were screened by study personnel for eligibility and enrolled on days 0 (date of birth) to 4 of age. Infants were not eligible while receiving parenteral antibiotics but could be enrolled if parenteral antibiotics were discontinued before day 4 of age. Detailed inclusion and exclusion criteria are listed in **Supplemental File, Section S2**.

The study was approved by the Hospital for Sick Children Research Ethics Board (REB #1000063899) and the ethical review committees at the International Centre for Diarrhoeal Disease Research, Bangladesh (icddr,b) (PR-19045) and Bangladesh Shishu Hospital and Institute (formally known as Dhaka Shishu Hospital), the ethical governing body for the Child Health Research Foundation (CHRF) (BICH-ERC-20/02/2019). Informed written consent was obtained from a parent or legal guardian before participant enrolment.

### Severe infection case definitions

Clinical severe infection (CSI) and possible CSI used in SEPSiS community-based surveillance:

*SEPSiS clinical severe infection (CSI)*: ≥1 of the following signs:

- Poor feeding (not sucking effectively or not sucking at all, on direct observation)
- Lethargy (movement only when stimulated or not moving at all, on direct observation)
- Convulsions (observed or strongly suspected by physician based on caregiver or community health research worker (CHRW) report)
- Severe chest in-drawing (observed)
- Fever (axillary temperature ≥37.5°C or rectal temperature ≥38°C)
- Hypothermia (axillary temperature <35.5°C or rectal temperature <36°C)

*Possible SEPSiS clinical severe infection (CSI)*: Equivalent to SEPSiS CSI with the addition of fast breathing (measured respiratory rate ≥60 breaths per minute) as a seventh sign.

Primary definition of severe infection:

At least one sign of SEPSiS clinical severe infection (CSI) documented by a study medical officer and/or non-study treating physician diagnosis of sepsis or another serious bacterial infection (SBI); AND at least one of the following two criteria:

- Non-study treating physician decision to admit to hospital, administration of ≥1 dose of a parenteral antibiotic on the day when SEPSiS CSI/sepsis/SBI is first ascertained, and treatment (or non-study treating physician intention to treat) with parenteral antibiotics for ≥5 consecutive days.
- Blood and/or cerebrospinal fluid (CSF) culture positive for a pathogenic bacterial or fungal organism.

Detailed explanations of the definition criteria are provided in **Supplemental File, Section S6**. A committee with expertise in infectious diseases and microbiology developed a list of blood pathogens and contaminants *a priori* and refined it after the study was concluded based on the isolation of organisms that were not pre-specified (**Supplemental File**, **Section S7**).

Variations in severe infection case definitions:

WHO clinical infection syndromes include pSBI, CSI and critical illness.^11^

*WHO possible serious bacterial infection (pSBI)*: ≥1 sign of pSBI (poor feeding, convulsions, severe chest indrawing, fever (≥38°C), hypothermia (<35.5°C), lethargy, or fast breathing (≥60 breaths per minute in infants <7 days old)) documented by a study physician and non-study physician decision to admit to hospital.

*Multiple signs of WHO pSBI*: ≥2 signs of pSBI documented by a study physician and non-study physician decision to admit to hospital.

*WHO clinical severe infection (CSI)*: ≥1 sign of CSI (poor feeding, severe chest indrawing, fever (≥38°C), hypothermia (<35.5°C), lethargy) documented by a study physician and non-study physician decision to admit to hospital. Note: WHO CSI (5 signs) is similar to SEPSiS CSI (6 signs) but excludes convulsions.

*WHO critical illness*: ≥1 sign of ‘critical illness’ (poor feeding, convulsions or lethargy) documented by a study physician and non-study physician decision to admit to hospital.

*Culture-confirmed severe infection*: Blood and/or cerebrospinal fluid (CSF) culture positive for a pathogenic bacterial or fungal organism and non-study physician decision to admit to hospital.

*Non-injury death*: any death that was not caused by an injury or accident per verbal autopsy.

A case definition was qualitatively considered to be *permissive* if it was expected to miss few cases of severe infection but might capture other diseases that were not severe bacterial infections (e.g., viral infections). A case definition was considered relatively *stringent* if it was expected to capture few non-severe infection diseases but could miss cases of severe infection.

### Community-based surveillance

Infants underwent surveillance for SEPSiS CSI from enrolment to 60 days of age. Up to 12 scheduled clinical assessments were performed by trained CHRWs. These assessments were conducted during in-person home or hospital visits or by telephone when an in-person visit was not feasible. At each scheduled visit, CHRWs assessed infants for signs of possible SEPSiS CSI. Infants with possible SEPSiS CSI ascertained by a CHRW were referred to the nearest study hospital for further assessment by a study medical officer. To identify illness episodes between scheduled visits, caregivers were asked to report any concerning signs to the study team via telephone. Infants with signs of possible SEPSiS CSI or other illness identified via an ad-hoc or follow-up assessment or phone call were referred to a study hospital for assessment by a study medical officer. Caregivers could also present directly to a study hospital to initiate an in-person visit with a study medical officer or a non-study treating physician, but also may have sought care at non-study facilities. The surveillance system to identify severe infection cases is shown in **Supplemental File, Figure S1.** At enrolment, study nurses also collected information on maternal and infant demographics.

### SEPSiS clinical severe infection and sepsis/serious bacterial infection case confirmation

When an infant with possible SEPSiS CSI presented to the study hospital, the study medical officer interviewed the caregiver and examined the infant. The infant was classified as having SEPSiS CSI if a study medical officer documented ≥1 sign of SEPSiS CSI. If the study medical officer documented SEPSiS CSI, the infant was referred to a non-study treating physician to confirm a clinical diagnosis of sepsis or SBI. A clinical diagnosis of sepsis was defined as sufficient general concern from the non-study treating physician to warrant a sepsis work-up (including blood culture). SBI was defined as an acute illness typically (or assumed to be) caused by bacteria. A list of infectious illnesses constituting SBI was established *a priori* and made available as a set of selectable response options for data entry (**Supplemental File, Section S3**). The study medical officer selected the suitable diagnostic label based on the non-study treating physician’s diagnosis. Diagnoses could also be manually entered by study personnel using free text. These free-text diagnoses were adjudicated by three physician members of the SEPSiS team and classified as ‘likely SBI,’ ‘possibly SBI,’ or ‘not SBI.’ Classification of manually entered ‘free text’ diagnoses is shown in **Supplemental File, Section S4**.

If an infant was admitted to a non-study facility, study medical officers attempted to obtain clinical and laboratory investigation data as soon as possible. In these cases, documentation of clinical signs of SEPSiS CSI by study personnel was not possible and the non-study treating physician diagnosis (as recorded by a study medical officer according to a standardized list of diagnostic labels) was used to determine a clinical diagnosis of sepsis or SBI.

### Sample collection and processing

A sepsis work-up was initiated if the infant was, or was intended to be, admitted to a hospital and had ≥1 sign of SEPSiS CSI present or had a diagnosis of sepsis and/or other SBI. Sepsis work-up laboratory analyses included complete blood count with differential, procalcitonin (PCT), high-sensitivity C-reactive protein (CRP), alanine aminotransferase (ALT), glucose, direct and indirect bilirubin, creatinine, aerobic blood culture, urine dipstick, urine microscopy and culture, real-time reverse transcriptase polymerase chain reaction (RT-PCR) on nasal swabs for influenza A and B, respiratory syncytial virus (RSV), ureaplasma and SARS-CoV-2, and skin swab for culture (where clinically indicated). If blood volume was lower than targeted (4.5 ml), assays were prioritized according to a pre-determined sequence (**Supplemental File, Table S1**). Although there was an intention to collect cerebrospinal fluid (CSF) where clinically indicated based on the non-study treating physician opinion, there were no instances in which CSF collection was performed.

For urine samples, clean catch or bagged urine collection was attempted. According to the operating procedures, if the urine dipstick was positive for leukocyte esterase and/or nitrites, a second sterile collection was to be attempted via bladder catheterization, but this was never performed. Mid-turbinate nasal swabs were collected by using a single swab to obtain epithelial cells from both nostrils. For some suspected skin or soft tissue infections, a skin swab was collected by gently rotating a sterile swab over the infected area, or a pus sample was obtained by needle aspiration. Details on sample processing are provided in **Supplemental File, Section S5**.

### Statistical analysis

A sample size of 2000 infants was estimated to obtain a desired precision of a 95% confidence interval (CI) width of 2%, assuming a baseline severe infection incidence proportion of 5% and loss to follow-up of 5%. Participant baseline characteristics were summarized using descriptive statistics and stratified by complete and incomplete follow-up. Frequencies and percentages were reported for categorical variables. Means, standard deviations, medians, 25^th^ and 75^th^ percentiles, minimum and maximum were computed for continuous variables. Incidence of severe infection, non-injury-related death, and variations in severe infection case definitions were estimated using incidence proportions and rates. Incidence proportions were calculated by plotting a Kaplan-Meier survival curve and subtracting the survival rate (proportion without the event) at 60 days from one. Incidence rates were calculated as the total number of severe infection episodes (using the primary and alternative severe infection definitions and accounting for multiple incident episodes per infant if these occurred) divided by the total number of infant-days at risk. CIs were generated using an intercept-only Poisson regression with robust standard errors to account for clustering within participants and a log offset of days at risk. Infants contributed person-time at risk if they were alive, had not voluntarily withdrawn from the study, were not hospitalized nor receiving parenteral antibiotics for a severe infection episode, had at least three severe infection-free days since the last day of administration of a prescribed parenteral antibiotic for an episode of severe infection or since the day of discharge from hospital (whichever was later), and were not lost to follow-up during the first 60 days of age. Loss to follow-up occurred if the infant missed the scheduled 3-month postnatal visit and the latest date of contact with study personnel was during the first 60 days of age. The date of loss to follow-up was defined as the latest date of contact with study personnel within the 60-day observation period post-birth. Detailed definitions of at-risk and not-at-risk periods are provided in **Supplemental File, Section S8**.

Proportions of severe infection cases identified from various pathways of the surveillance system were estimated. Laboratory investigation results and bacterial and viral etiologies for cases of severe infection were also summarized.

Additional analyses explored the effect of variations in severe infection case definitions on incidence estimates within subgroups, including gestational age at enrolment (<37 weeks and ≥37 weeks), age at onset of severe infection (<28 days and 28-60 days), and sex. Sensitivity analyses for the calculation of the primary severe infection definition incidence proportions and rates were performed and included: (i) restricting to infants with ≥30 days of follow-up time; (ii) varying the categorization of manually entered free-text SBI diagnoses; (iii) varying the duration of the severe infection-free period to determine re-entry into the subsequent at-risk period; (iv) restricting the observation period to 0-59 days; and (v) using the last scheduled date of contact with study personnel to define the date of loss to follow-up. Rationales for these analyses are described in **Supplemental File, Section S9**.

Information on independent verification and code review for analyses is provided in **Supplemental File, Table S2**. All statistical analyses were conducted using R version 4.2.3.

### Patient and public involvement

Caregivers/public were not involved in developing the research question, recruitment, design and conduct of the study, or methods for study result dissemination.

## RESULTS

Of 1939 enrolled infants (**Figure 1** and **Table 1**), 1522 (78%) had complete follow-up and 417 (22%) had incomplete follow-up (left catchment area at least once and/or lost to follow-up). There were no substantial differences in maternal and infant characteristics between infants who had complete follow-up compared to those who had incomplete follow-up (**Table 1**).

**Figure 1.**
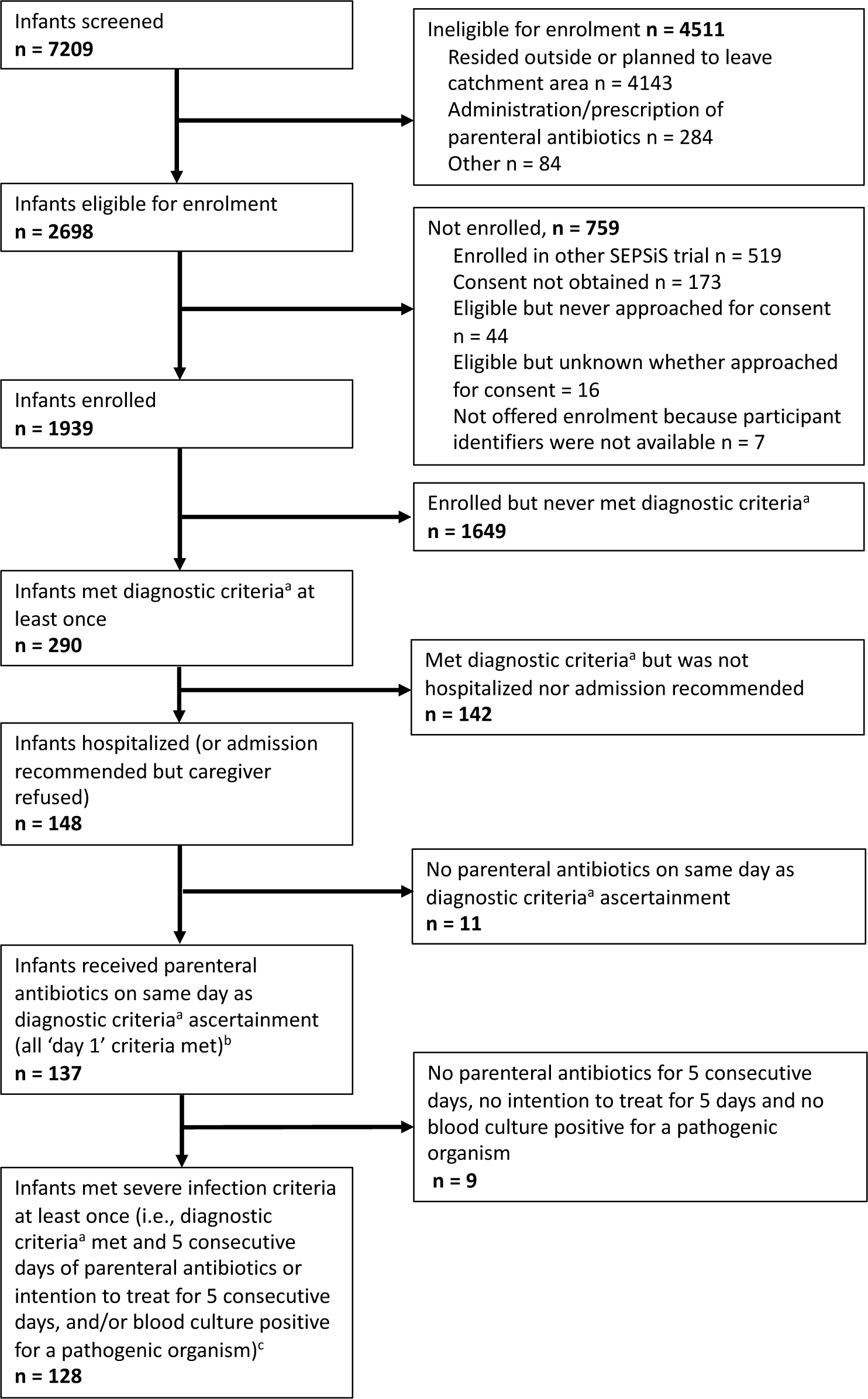
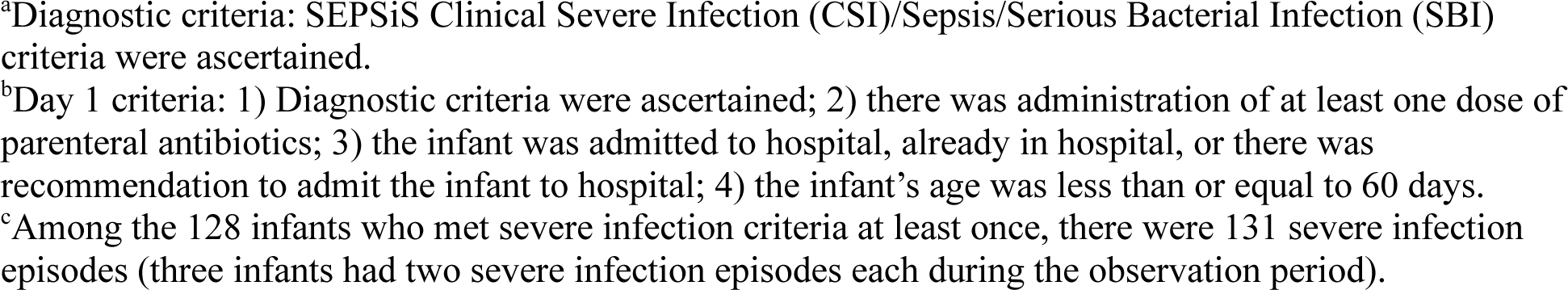
Study flow diagram.

**Table 1.**
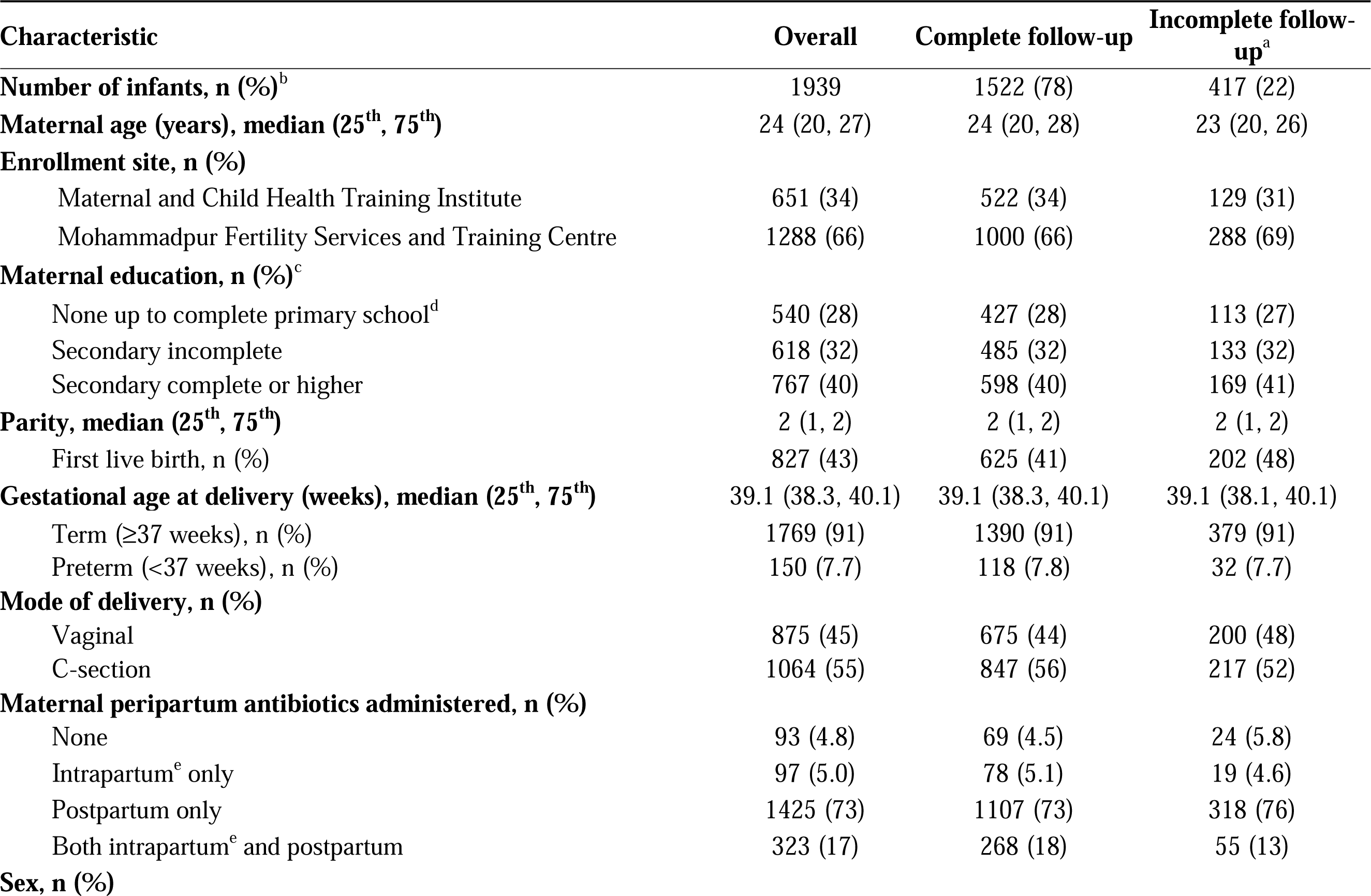
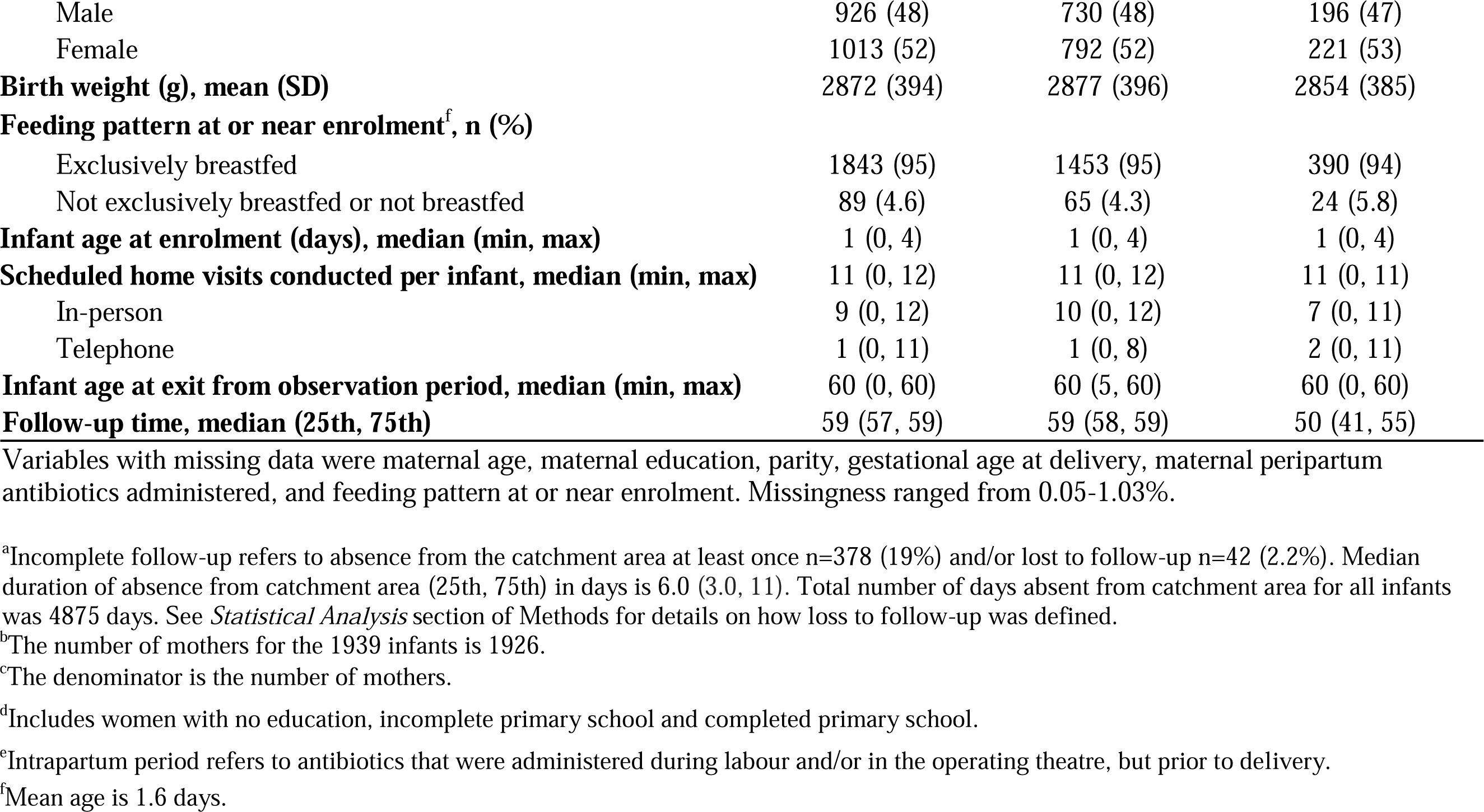
Participant characteristics, overall and by complete versus incomplete follow-up.

Using the primary severe infection definition, there were 131 severe infection episodes among 128 infants during the first 60 days of age (**Figure 1 and Supplemental File, Figure S2**). Three infants each had two incident severe infection episodes during the first 60 days of age (**Table 2**). The severe infection incidence rate per 1000 infant-days at risk was 1.2 (0.97-1.4) using the primary definition and 0.84 (0.69-1.0) using WHO pSBI. Given that WHO CSI, WHO critical illness and multiple signs of WHO pSBI are subsets of WHO pSBI, they each had lower incidence rates than WHO pSBI (**Table 2** and **Supplemental File, Figure S3**). The differences in incidence rates by case definition were maintained within subgroups including gestational age at enrolment (<37 weeks and ≥37 weeks), age at onset of severe infection (<28 days and 28-60 days of age), and sex (**Supplemental File, Table S3, Table S4 and Table S5**). Three infants had a severe infection episode with a positive blood culture for a pathogenic organism. Seven infants died from non-injury-related causes. Of these seven deaths, two infants had a severe infection leading to their death. The incidence rate per 1000 infant-days at risk of blood culture-confirmed severe infection was 0.026 (0.0085-0.081), while that of non-injury death was 0.061 (0.029-0.13). The incidence rate of severe infection using the primary definition and non-injury deaths was 1.2 (1.0-1.4) per 1000 infant-days at risk. Time from birth to severe infection for the primary and alternative definitions is illustrated in **Figure 2**.

**Figure 2.**
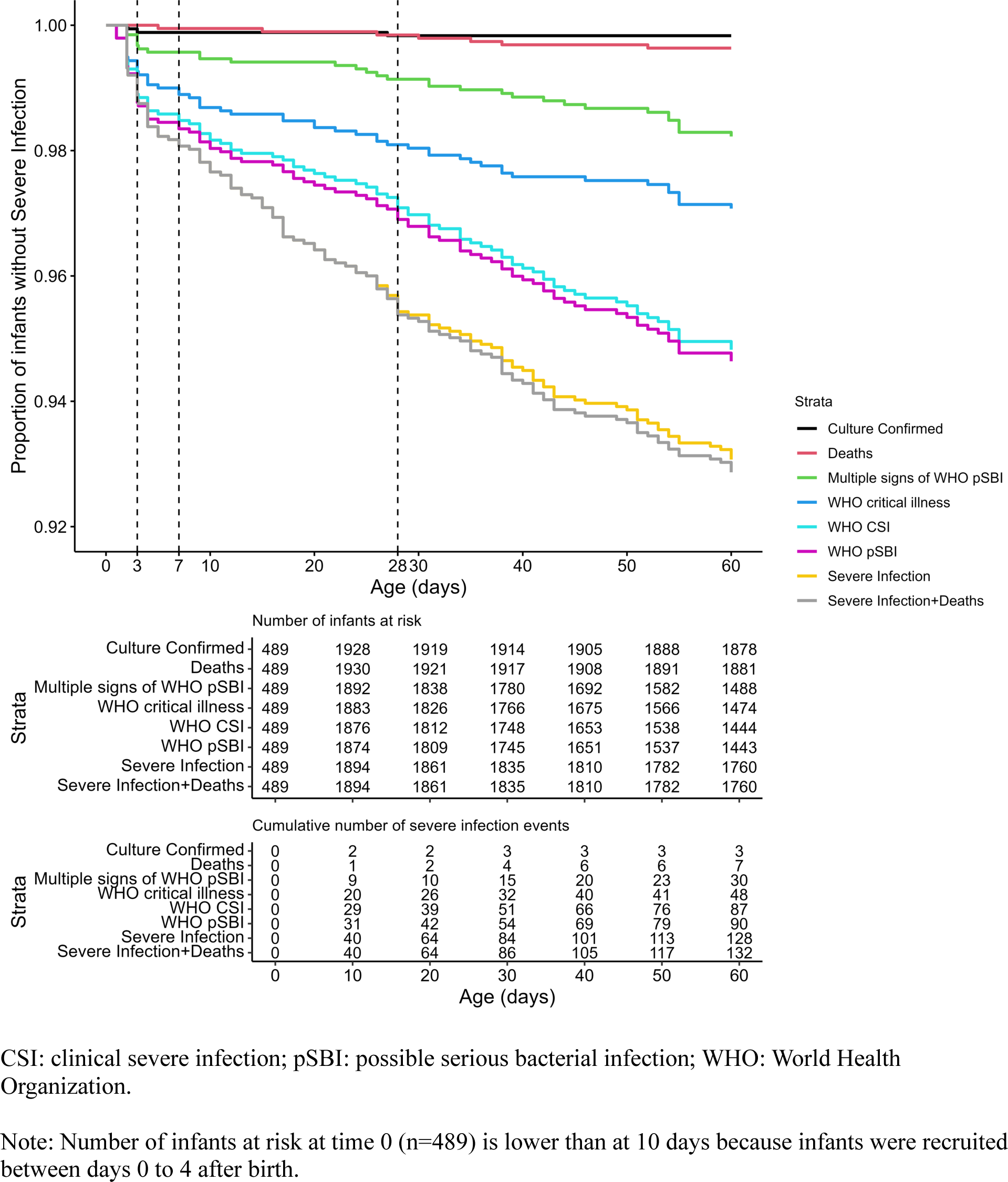
Kaplan-Meier curves for time to severe infection up to 60 days of age according to various severe infection case definitions.

**Table 2.** Incidence proportions and incidence rates of severe infection and variations in severe infection case definition (N_infants_=1939).

| **Severe infection case definition** | **Number of events** | **Number of infants with at least one event** | **Infant-days at risk** | **Incidence proportion<sup>c</sup>, per 1000 infants at risk (95% CI)** | **Incidence rate, per 1000 infant-days at risk (95% CI)** |
| --- | --- | --- | --- | --- | --- |
| **Severe infection** | 131 | 128 | 113238 | 69 (58, 81) | 1.2 (0.97, 1.4) |
| **WHO possible serious bacterial infection<sup>a</sup>** | 92 | 90 | 109316 | 54 (42, 65) | 0.84 (0.69, 1.0) |
| WHO clinical severe infection<sup>b</sup> | 88 | 87 | 109327 | 52 (41, 63) | 0.80 (0.65, 0.99) |
| WHO critical illness<sup>c</sup> | 48 | 48 | 109677 | 29 (21, 38) | 0.44 (0.33, 0.58) |
| Multiple signs of WHO possible serious bacterial infection | 30 | 30 | 109838 | 18 (11, 24) | 0.27 (0.19, 0.39) |
| **Culture-confirmed severe infection** | 3 | 3 | 114335 | 1.7 (0.0, 3.6) | 0.026 (0.0085, 0.081) |
| **Death (non-injury)** | 7 | 7 | 114384 | 3.6 (0.95, 6.3) | 0.061 (0.029, 0.13) |
| **Severe infection + Deaths (non-injury)<sup>d</sup>** | 136 | 132 | 113238 | 69 (57, 80) | 1.2 (1.0, 1.4) |

WHO: World Health Organization.
<sup>a</sup>At least one sign of possible serious bacterial infection (poor feeding, convulsions, severe chest indrawing, fever ( $\geq 38^{\circ}\text{C}$ ), hypothermia ( $< 35.5^{\circ}\text{C}$ ), lethargy, or fast breathing ( $\geq 60$ breaths per minute in infants $< 7$ days old)) documented by a study medical officer and non-study treating physician decision to admit to hospital.
<sup>b</sup>At least one sign of clinical severe infection (poor feeding, severe chest indrawing, fever ( $\geq 38^{\circ}\text{C}$ ), hypothermia ( $< 35.5^{\circ}\text{C}$ ), or lethargy) documented by a study medical officer and non-study treating physician decision to admit to hospital.
<sup>c</sup>At least one sign of critical illness (poor feeding, convulsions, or lethargy) documented by a study medical officer and non-study treating physician decision to admit to hospital.
<sup>d</sup>Not a cumulative sum of severe infection and deaths because 2 infants had severe infection and then died.
<sup>e</sup>Estimated as 1 minus the survival probability at day 60. [Note: the analysis plan in the SEPSiS observational cohort study protocol also specified an additional method to calculate incidence proportions (dividing the number of severe infection events up to 60 days of age by the number of infants enrolled); however, we found the methods generated similar estimates, and we only reported the results using the Kaplan-Meier method because it accounts for right-censoring of infants (due to loss to follow-up or death) and was therefore considered to be less biased].

In all sensitivity analyses, there were no substantial differences in severe infection incidence proportions or rates using the primary definition (**Supplemental File, Table S6**).

Of 131 primary definition severe infection episodes, the proportion with a completed sepsis work-up laboratory test ranged from 59% to 85%, depending on the test (**Supplemental File, Table S7**). Of severe infection episodes for which sepsis work-up tests were done, the proportion of test results meeting their respective thresholds for clinical concern ranged from 4.8% for elevated ALT to 24% for elevated CRP. In the cohort, there were three positive blood cultures for a pathogenic organism, eight positive urine cultures for a pathogenic organism and four positive skin swab cultures taken from sites with suspected infections (**Supplemental File, Table S8**). The primary severe infection definition captured all these events involving positive cultures, whereas the WHO pSBI definition only captured two of the three positive blood cultures, six of the eight positive urine cultures, and none of the four positive skin swab cultures. Of primary severe infection definition episodes, 51% had a RT-PCR nasal swab, at least one of white blood cell count, neutrophil count, CRP or PCT, and a blood culture completed (**Supplemental File, Table S9**). Of severe infection episodes for which these tests were done, 70% had a probable viral and/or bacterial infection (RT-PCR nasal swab positive for either RSV/influenza, evidence of a systemic inflammatory response, or a positive urine dipstick or urine or blood culture). Most (67%) episodes were probable viral illnesses without bacterial infection (negative urine dipstick and bacterial cultures without evidence of a systemic inflammatory response, or RSV/influenza positive). Only 36% were possible or confirmed bacterial infections without evidence of viral etiology (evidence of a systemic inflammatory response and/or a positive urine dipstick or bacterial culture, and RSV/influenza negative).

Of the 131 severe infection episodes using the primary definition, 19 (15%) were identified directly following a scheduled CHRW home visit assessment and CHRW referral, and 112 (85%) were identified following caregiver self-referral (**Figure 3**). For 43 (33%) severe infection episodes, the infants were assessed by a non-study treating physician without prior assessment by study personnel and met criteria for the primary severe infection definition through a non-study treating physician diagnosis of sepsis/SBI rather than documentation of clinical signs of SEPSiS CSI.

**Figure 3.**
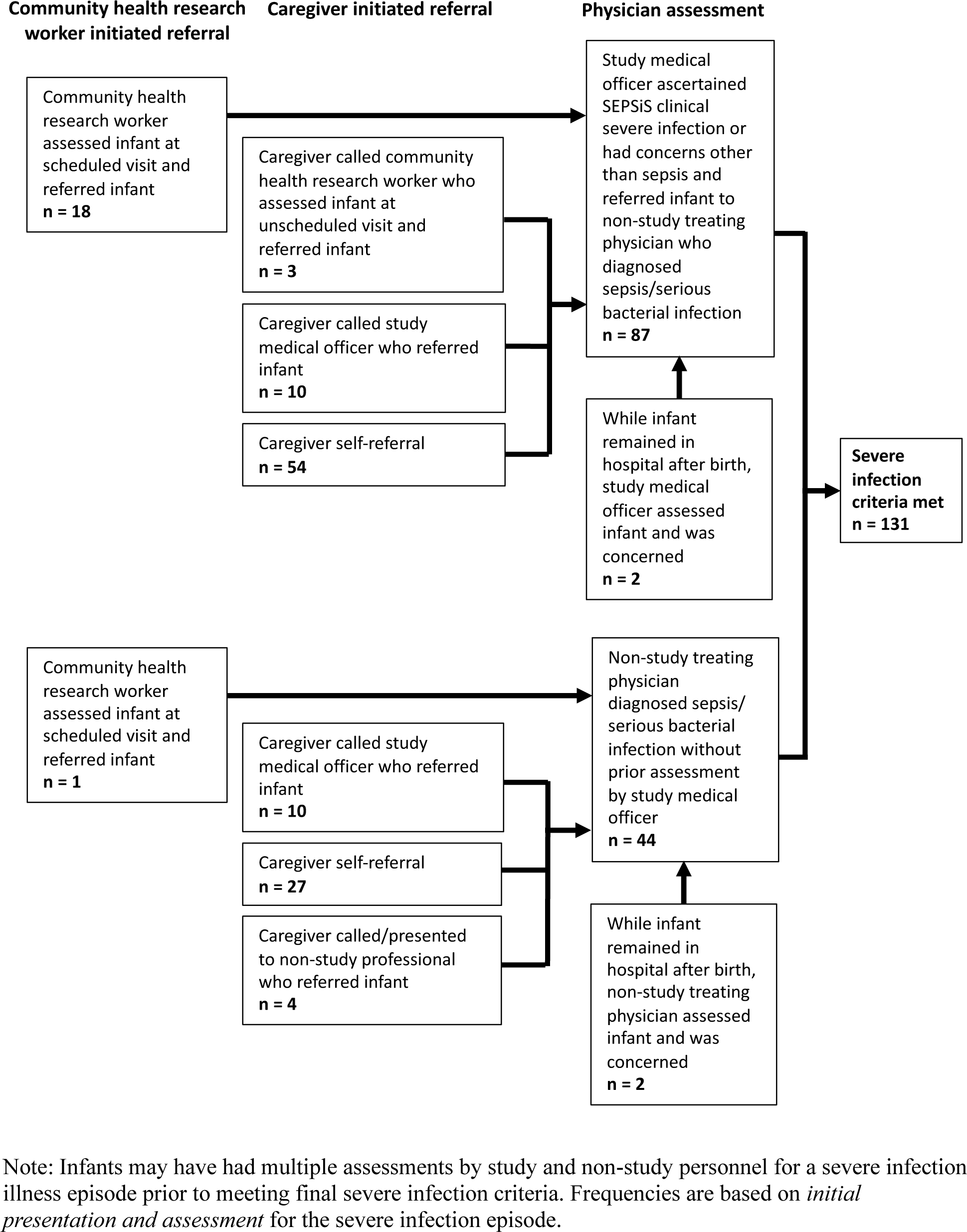
Referral pathways by which severe infection episodes were identified within the study.

We conducted sample size calculations for a theoretical severe infection prevention RCT for young infants using various case definitions of severe infection (**Supplemental File, Table S10**). The number of infants required per trial group is lowest using the primary severe infection definition (which had the highest incidence) and increases to very high sample sizes when using culture-confirmed infection.

## DISCUSSION

The incidence of severe infection up to 60 days of age in Bangladeshi infants born generally healthy varied considerably by case definition. WHO clinical sign-based definitions such as pSBI yielded lower incidence rates than the primary severe infection definition, which comprised a more complex set of therapeutic and microbiologic criteria including either the administration of ≥5 consecutive days of parenteral antibiotics and/or blood-culture confirmation. In practice, the primary definition was more permissive because the diagnostic component of the definition could be fulfilled by either a clinical sign documented by a study medical officer or a non-study treating physician diagnosis of sepsis or SBI, which enabled the definition to be applied to episodes for which a standardized assessment of clinical signs performed by study personnel was absent. Because many infants presented for care via self-referral to non-study physicians, definitions that required documentation of specific clinical signs by study personnel led to a substantial proportion of cases of non-study physician-diagnosed sepsis/SBI being missed.

The majority (85%) of severe infection cases using the primary definition were identified following caregiver self-referral, and it is plausible that many of the other cases identified following a CHRW scheduled visit may have also eventually been identified following caregiver self-referral, had the CHRW visit not been scheduled. In an urban setting such as Dhaka, health facilities are numerous and caregivers may thus seek care early for infant illness. Therefore, when conducting young infant severe infection prevention RCTs in such settings, scheduled home assessments by study personnel to identify infants requiring referral may not be warranted, and resources may be better allocated toward other operational aspects of the RCT, such as staffing of study personnel at study hospitals and implementing mechanisms to retroactively identify cases that present to non-study physicians. However, in settings where caregivers have limited access to health facilities and may not readily seek care for infant illness (e.g., some rural settings), or infants are receiving treatment including antibiotics at home which may affect the yield of subsequent investigations (e.g., blood culture), more frequent scheduled home assessments by study personnel to identify infants requiring referral may be warranted. It is also possible that frequent scheduled home visits could have facilitated earlier identification and management of mild infections, thereby preventing their progression to severe infections. This may affect incidence estimates and warrants consideration in RCT design.

In a previous observational study investigating the causes and incidence of community-acquired serious infections among infants 0-60 days of age in Bangladesh, India and Pakistan (ANISA), the incidence per 1000 live births was 95 for WHO pSBI and 1.6 for culture-confirmed bacterial infection.^13^ Notably, the SEPSiS observational study cohort was not a formal birth cohort and during the enrolment period from days 0 to 4 of age, infants were not eligible to be enrolled while receiving parenteral antibiotics. This likely led to the exclusion of cases of early-onset sepsis, which was intentional in the design of the SEPSiS study since its purpose was to guide the design of RCTs for severe infection prevention by postnatal interventions rather than treatment or prenatal or perinatal prevention. Therefore, it was expected that the incidence proportion of WHO pSBI in the SEPSiS cohort would be lower than in the ANISA cohort. However, the incidence proportion of culture-confirmed cases in the SEPSiS cohort was similar to the ANISA cohort. This similarity in the incidence of culture-confirmed cases was unexpected and may have been due to differences in blood culture processing techniques, organisms considered to be pathogenic, and infectious disease specialist classification of isolates. In a cohort of facility-born infants in seven LMICs across South Asia and Africa (BARNARDS), the incidence estimates of sepsis among infants 0-60 days of age were substantially higher than the estimates in both the SEPSiS and ANISA cohorts with 166 cases of clinically suspected sepsis per 1000 live births and 46.9 cases of blood culture-confirmed sepsis per 1000 live births.^4^ In addition to the SEPSiS cohort likely excluding many cases of early-onset sepsis, possible reasons for these discrepancies include differences in definition criteria and different *a priori* determination of pathogenic organisms and post-hoc classification of isolates. These discrepancies further highlight that there may be substantial differences in incidence estimates of severe infection in young infants depending on the case definition used and source populations from which cases arise.

When formulating a case definition for young infant severe infection in LMICs, important considerations include balancing permissiveness and stringency and resources available to operationalize the case definition. The primary severe infection definition in the SEPSiS study was permissive by allowing for inclusion of cases that met clinical sign criteria and/or physician diagnosis of sepsis/SBI, but stringency was imposed by requiring administration of parenteral antibiotics for ≥5 days and/or blood culture-confirmation. The criteria of treatment or intention to treat with parenteral antibiotics for ≥5 days objectively indicates a high level of clinical concern from a physician, and has been used in case definitions in infant sepsis prevention RCTs.^14^ Compared to the WHO pSBI definition, the primary severe infection definition had higher sensitivity for capturing positive blood and urine cultures for pathogenic organisms. However, the laboratory investigation results of primary severe infection episodes suggest that this definition is likely still non-specific, since only about one third of severe infection episodes were characterized as possible or confirmed bacterial infections based on laboratory criteria (i.e., systemic inflammatory response and/or a positive urine dipstick or bacterial culture, and RSV/influenza negative). While ∼30% of episodes may have been non-infectious given they had no evidence of a systemic inflammatory response, were RSV/influenza negative, and had negative urine dipstick and bacterial cultures, some of these episodes may have been caused by a non-RSV/influenza virus. The small proportion of blood culture positive cases was partly due to challenges with adherence to testing protocols (e.g., obtaining samples prior to antibiotic administration), but also because many cases meeting severe infection criteria were probably not bacterial infections.

International consensus criteria that incorporate laboratory investigation results, such as the Phoenix Sepsis Score, have been developed to identify sepsis in infants and children 0-18 years of age, and are intended to be globally applicable.^15^ However, in the present study, the intended sepsis work-up laboratory investigations were only obtained in about half of severe infection episodes using the primary definition. Therefore, operationalization of international consensus criteria such as the Phoenix Sepsis Score may not be feasible in similar LMIC settings. A consensus definition of severe infection in young infants that balances permissiveness and stringency and can be operationalized in LMICs would align research in this area and improve comparability of RCTs.

The SEPSiS observational cohort study was conducted over the course of the COVID-19 pandemic.^16^ In young infants, SARS-CoV-2 generally causes a mild viral illness without major acute complications^17,18^ and only one infant with severe infection using the primary definition was SARS-CoV-2 positive. Therefore, it is unlikely that SARS-CoV-2 directly affected incidence estimates of severe infection in this study. However, it is possible that the COVID-19 pandemic may have indirectly affected severe infection incidence estimates in both directions. For example, care-seeking behaviour may have decreased or been delayed during lockdown periods, leading to higher severe infection incidence.^19^ Conversely, social distancing measures may have decreased pathogen exposure leading to lower severe infection incidence. It is difficult to predict the overall effect of the pandemic on young infant severe infection incidence estimates, but the pandemic is unlikely to have affected the differences in incidence estimates by case definition found in this study.

This study had several limitations. First, given that during the enrolment period from days 0 to 4 of age, infants were not eligible while receiving parenteral antibiotics, the incidence estimates of this study do not represent those of a birth cohort. However, the intention of this study was to inform the design of RCTs of severe infection prevention interventions in the postnatal period. Trialists can therefore use these findings to inform the selection of their case definition, sample size calculations and design of surveillance systems to identify cases. Second, the intended sepsis work-up laboratory investigations were only obtained in about half of severe infection cases using the primary definition, such that the inferences based on the laboratory investigation results may not be generalizable to all severe infection episodes. Lastly, the RT-PCR nasal swabs were limited to detection of RSV, influenza, SARS-CoV-2 and ureaplasma. More extensive microbiologic panels may have better characterized the causes of severe infection episodes.

## CONCLUSION

In an observational cohort study in Dhaka, Bangladesh, the incidence of severe infection in young infants varied considerably by case definition. A severe infection definition that requires physician documentation of standardized clinical signs may miss a substantial proportion of cases identified by physician diagnosis of sepsis/SBI. A consensus definition of severe infection in young infants that balances permissiveness and stringency and can be operationalized in LMICs would improve the comparability of RCTs. If health facilities are accessible and caregivers seek care for infant illness, frequently scheduled home assessments by study personnel to identify infants requiring referral may not be warranted.

## DATA AVAILABILITY STATEMENT

De-identified datasets and code files used in the analyses of this study are publicly available at the Borealis online data repository at https://borealisdata.ca/dataset.xhtml?persistentId=doi:10.5683/SP3/MOSXFC under Custom Dataset Terms. The statistical analysis plan (SAP) for this study that specifies the definition and procedures for the derivation of the ‘severe infection’ variable is available at https://borealisdata.ca/dataset.xhtml?persistentId=doi:10.5683/SP3/JEDIJY. Standard operating procedures (SOPs), including those related to SEPSiS observational cohort study enrolment, consent and data collection procedures, are available at https://borealisdata.ca/dataset.xhtml?persistentId=doi:10.5683/SP3/WKDQYY.

## FOOTNOTES

### Contributors

AF, CH, LGP, DGB, PSS, SKM, DHH, MSI, AAM, EP, SKS, RH, MIH, AE, KMOC, MGL, SS (Shamima Sultana), MMH, TA, SAS and DER conceptualized the study and developed the methodology. DER was the principal investigator. SAS led the project investigation and data collection at the International Centre for Diarrhoeal Disease Research, Bangladesh site. SKS led the project investigation and data collection at the Child Health Research Foundation site. DER led the project investigation at the Hospital for Sick Children site. MSI, AAM, SKS, RH, MIH, SS (Shamima Sultana), SMMB, SMAG, EK, SS (Sharika Sayed), SY, MMH and SAS contributed to data generation and collection and supervision of field or lab activities. AF, CH, C-YC and AE conducted the formal analysis of the data. DGB and DER supervised data analysis. AF wrote the initial draft of the manuscript. All authors provided feedback on data analysis and the manuscript. All authors read and approved the final manuscript. Funding acquisition and project administration were led by DER. DER takes full responsibility for the integrity of the work as a whole from inception to the published paper.

### Patient and public involvement

Caregivers/public were not involved in developing the research question, recruitment, design and conduct of the study, or methods for study result dissemination.

## Supporting information

STROBE Checklist

## Acknowledgements

We thank Katarina Vukovojac and Aline Freitas for contributing some of the statistical code that was used in or adapted for this analysis. We also thank all SEPSiS co-investigators and study personnel who contributed to the SEPSiS observational cohort study.

## Ethics approval

This study was approved by the Hospital for Sick Children Research Ethics Board (REB #1000063899) and the ethical review committees at the International Centre for Diarrhoeal Disease Research, Bangladesh (icddr,b) (PR-19045) and Bangladesh Shishu Hospital and Institute (formally known as Dhaka Shishu Hospital), the ethical governing body for the Child Health Research Foundation (CHRF) (BICH-ERC-20/02/2019). Informed written consent was obtained from a parent or legal guardian before participant enrolment.

