## Supplemental File for "Severe infection among young infants in Dhaka, Bangladesh: effect of case definition on incidence estimates"

**Table of contents**	Page

**Section S1**. Additional details of study sites	2 **Section S2**. Inclusion and exclusion criteria	2

**Figure S1.** Surveillance system to identify severe infection cases	4

**Section S3**. *A priori* list of infectious illnesses that constitute a serious bacterial 5 infection (SBI)

**Section S4**. Classification of manually entered ‘free text’ diagnoses	5

**Table S1**. Prioritization of blood collected from sepsis work-up	6

**Section S5**. Sample processing	6

**Section S6.** Primary severe infection definition criteria	7

**Section S7.** Categorization of organisms in blood as pathogens or contaminants  8

**Section S8**. At-risk period definition	9

**Section S9.** Sensitivity analyses	10

**Table S2.** Summary of analyses that were independently verified and reviewed	11

**Figure S2**. Flow diagram from diagnostic criteria to severe infection criteria,	12

within the first 60 days after birth, in infant days

**Figure S3**. Coefficient plot comparing incidence rate estimates of severe infection and	14

variations in severe infection case definitions.

**Table S3.** Incidence proportions and incidence rates of severe infection by gestational age	15

**Table S4.** Incidence proportions and incidence rates of severe infection by age at onset of	16

event

**Table S5.** Incidence proportions and incidence rates of severe infection by sex	17

**Table S6**. Sensitivity analyses for primary severe infection case definition incidence	18

proportion and rate

**Table S7**. Laboratory investigation results	20

**Table S8.** Positive cultures captured by the primary *severe infection* definition and the	23

WHO *possible serious bacterial infection* (pSBI) definition

**Table S9**. Characterization of severe infection episodes by RSV/influenza positivity, 24 biomarkers, urine dipstick results and urine and blood culture positivity

**Table S10**. Sample size calculations for a theoretical randomized controlled trial of a severe	26

infection prevention intervention in young infants

**Section S1.** Additional details of study sites

Located in the Azimpur area of Dhaka city, Maternal and Child Health Training Institute (MCHTI) is a public secondary-level facility that provides free or low-cost health care to pregnant women and children. At the time the study was initiated, the labour and delivery unit at MCHTI had 152 beds and performed an average of 360 deliveries per month. Mohammadpur Fertility Services and Training Centre (MFSTC) is located in the Mohammadpur area of Dhaka city, approximately 7 kilometres from MCHTI. When the study began, the labour and delivery unit at MFSTC had 72 beds and performed an average of 442 deliveries per month. Both MCHTI and MFSTC are staffed with pediatricians, obstetricians, nurses, paramedics and other allied health professionals.

**Section S2.** Inclusion and exclusion criteria

*Inclusion criteria*

1. Infants up to and including 4 days of age

2. Infant delivered at a study hospital

3. Orally feeding currently^a^

4. Informed consent by parent or guardian

5. Intends to maintain residence within the defined catchment areas (upon discharge from hospital) until 60 days of age

*Exclusion criteria*

1. Birth weight <1500g^b^

2. Death or major surgery^c^ considered to be highly probable within the first week of life

3. Major congenital anomaly of the gastrointestinal tract

4. Maternal HIV infection and/or history of mother ever receiving anti-retroviral drug(s) for presumed HIV infection^d^

5. Current mechanical ventilation and/or cardiac support (e.g., inotropes) and/or administration/prescription of parenteral antibiotics^a^

6. Any prenatal or postpartum use of non-dietary probiotic supplement by the mother (during current pregnancy)^e^

7. Any postnatal administration of non-dietary probiotic or prebiotic supplements to infant

8. Enrolment of infant in any other clinical trial involving the administration of probiotics and/or prebiotics

9. Resides in the same household as another infant previously enrolled in the study, or any study within the research platform, who is currently <60 days of age; however, twins may be enrolled simultaneously

10. Multiple gestation for which the number of liveborn infants from the same pregnancy exceeds two (i.e., triplets or higher order multiples)

^a^These criteria were time-varying, so they were reassessed on a daily basis until no longer eligible for another reason (i.e., beyond day 4 of life), as long as the infant remained potentially eligible by other criteria. Orally feeding was defined as being able to take a probiotic or synbiotic supplement by mouth on a daily basis.

^b^Current infant weight, as measured and documented by study personnel, was used if birth weight was missing, illegibly recorded, or suspected of being an error (e.g., implausible value, discrepancy of greater than 15% between documented birth weight and measured current weight).

^c^Major surgery was defined as an operative procedure to explore and/or repair an organ or tissue that is performed under general anesthesia. Examples relevant to the neonatal period include ligation of patent ductus arteriosus, repair of abdominal wall defects, repair of bowel perforation due to necrotizing enterocolitis, repair of tracheoesophageal fistula and/or esophageal atresia, and repair of myelomeningocele. Conversely, examples of common procedures in newborns not considered major surgery include circumcision, tongue tie release, and removal of extra digits (polydactyly).

^d^Although HIV in very young infants was expected to be rare in this context, there was a low but non-zero theoretical risk that a baby born to an HIV-positive mother could be significantly compromised, particularly in cases where in-utero transmission occurred earlier in pregnancy. Both the HIV-positive mother and infant were expected to represent a unique population in regard to their respective microbiomes and excluding them was not expected to affect the generalizability of results to the population as a whole. The number of HIV-positive infants was anticipated to be too low to conduct sub-group analyses, and thus, it would not be possible to make meaningful inferences about this population, even if they were to be included in the study.

^e^A non-dietary probiotic is a commercial (store-bought) probiotic product that is consumed in the form of a capsule, powder, liquid, etc., although it may be mixed into a food or drink at the time of consumption. In contrast, a dietary probiotic is an ingredient of a food or beverage that either occurs naturally or is added during home production or the commercial manufacturing process (e.g., yogurt or fermented drinks).

**Figure S1.** Surveillance system to identify severe infection cases

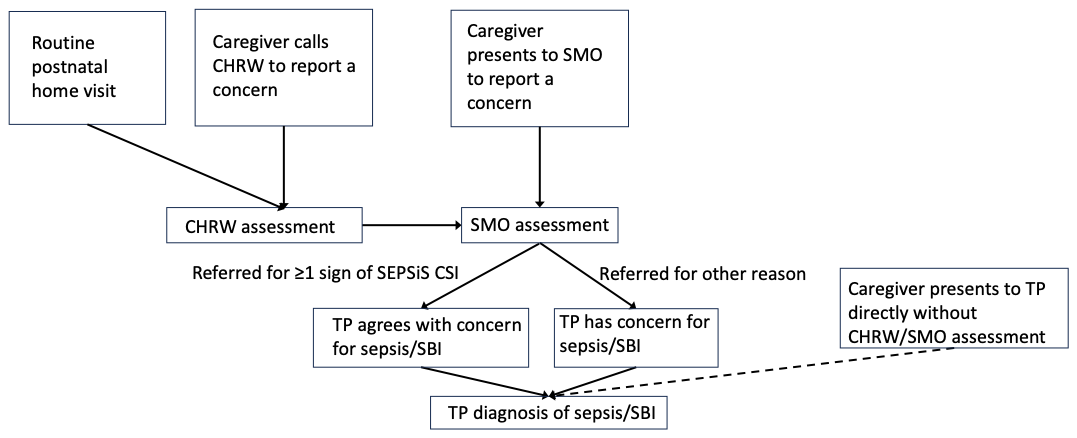

CHRW: Community health research worker (study personnel)

SMO: Study medical officer (study personnel)

TP: Non-study treating physician

CHRW/SMO referral pathway: Infant was assessed by a SMO with or without CHRW assessment prior to SMO assessment. SMO ascertained ≥1 SEPSiS CSI sign and then referred the infant to a TP for concern for sepsis/SBI, or the SMO did not ascertain ≥1 SEPSiS CSI sign and then referred the infant to a TP for another reason (i.e., not for signs of SEPSiS CSI).

Direct to TP pathway: Infant likely was not assessed by a CHRW or SMO

and caregiver presented directly to a hospital and was assessed by a TP

**Section S3.** *A priori* list of infectious illnesses that constitute a serious bacterial infection (SBI)

Meningitis, bacteremia/septicemia, pneumonia, urinary tract infection, osteomyelitis, septic arthritis, purulent conjunctivitis, omphalitis, pyogenic soft tissue infections (unusual skin rash, lesions or abscesses that are not ‘dry’), dysentery.

**Section S4.** Classification of manually entered ‘free text’ diagnoses

Classification of manually entered ‘free text’ diagnoses (variations in spelling were permitted but are not shown in this table).

| **Serious bacterial infection label** | **Free text labels considered ‘possibly serious bacterial infection’ (permissive)** | **Free text labels considered ‘likely serious bacterial infection’ (stringent)** |
| --- | --- | --- |
| Suspected sepsis | NA | Septicaemia, NEC (necrotizing enterocolitis), neonatal sepsis |
| Pneumonia | Right pulmonary inflammatory lesion | Aspiration pneumonia, bronchopneumonia, pneumonia, suspected pneumonia, congenital pneumonia |
| Fever without a localizing sign | Fever | NA |
| Suspected meningitis | NA | Suspected meningitis, meningitis, meningoencephalitis |
| Omphalitis | Discharging umbilicus, umbilical oozing, umbilical redness | Infection of umbilical stump, umbilical sepsis, umbilical infection, suspected umbilical sepsis |
| Urinary tract infection | Suspected UTI | UTI, urinary tract infection, Lower UTI with labia fusion |
| Purulent conjunctivitis | Conjunctivitis, eye discharge | Eye infection |
| Pyogenic soft tissue infection | Skin infection, tubercular abscess, soft tissue infection, skin pustule | Axillary boil, boil |
| Dysentery | Invasive diarrhea, persistent diarrhea, amoebic dysentery | NA |
| Osteomyelitis, Septic arthritis | NA | NA |

**Table S1.** Prioritization of blood collected from sepsis work-up^a^

| **Blood volume drawn (ml)** | **BACTEC bottle (ml)** | **EDTA Tube #1 – hematology (ml)** | **EDTA tube #2 – molecular (ml)** | **EDTA tube #3 – plasma (ml)** | **Red Top Tube – biochemistry + archive (ml)** | **Whatman paper filter card^b^ (μl)** |
| --- | --- | --- | --- | --- | --- | --- |
| **4.5** | 1 | 0.5 | 0.5 | 0.5 | 2 | ~50 |
| **>4 – < 4.5** | 1 | 0.5 | 0.5 | Remaining | 2 | ~50 |
| **>3 – ≤4** | 1 | 0.5 | Nil | Nil | Remaining (1.5 – <2.5 ml) | ~50 |
| **>2 – ≤3** | 1 | 0.5 | Nil | Nil | Remaining (0.5 – <1.5 ml) | ~50 |
| **>1.5 – ≤2** | 1 | Nil | Nil | Nil | Remaining (0.5 – <1.0 ml) | ~50 |
| **>1 – ≤1.5** | 1 | ~0.5 | Nil | Nil | Nil | ~50 |
| **≤1** | Whole amount | Nil | Nil | Nil | Nil | Any remaining |

^a^This chart served as a general guideline; however, in practice, specific volumes may have varied by patient. Also, if requested by the non-study treating physician, prioritization of aliquots may have been altered to address the clinical requirements.

^b^If possible, excess blood in the collection tubing (up to approximately 50 µl) was spotted onto a Whatman filter paper card and dried for at least 3 hours at room temperature. Dried blood spot cards were archived at -70°C or colder for future processing.

**Section S5.** Sample processing

Blood samples were aerobically cultured in the BACTEC 9120 blood culture instrument as soon as possible after collection. If the bottle was beep-positive for bacterial growth or was suspected to be positive, the broth was sub-cultured on solid medium and checked for Gram-staining to identify pathogenic bacteria. A primary culture was performed for BACTEC bottles before incubation if the transfer of samples to the culture instrument was delayed by more than 8 hours after collection. Urine samples were sub-cultured on selective media to isolate and identify pathogenic bacteria. Nucleic acids were extracted from nasal swabs using commercially available nucleic acid isolation kits and analyzed using a one-step singleplex RT-PCR assay for detection and identification of influenza A and B, respiratory syncytial virus (RSV), and ureaplasma. Additional viral or bacterial pathogens, including SARS-CoV-2, were added to the qPCR panel, depending on technical feasibility and available funding. Real-time RT-PCR reactions were carried out using primers and corresponding fluorescent probes optimized to specific targets on respiratory pathogens. Skin swabs were processed immediately upon arrival at CHRF and cultured on a solid medium.

**Section S6.** Primary severe infection definition criteria

Primary definition of severe infection

At least one sign of SEPSiS clinical severe infection (CSI) documented by a study medical officer and/or non-study treating physician diagnosis of sepsis or another serious bacterial infection (SBI)^a^; AND at least one of the following two criteria:

Non-study treating physician decision to admit to hospital, administration of ≥1 dose of a parenteral antibiotic on the day when SEPSiS CSI/sepsis/SBI is first ascertained, and treatment (or non-study treating physician intention to treat) with parenteral antibiotics ≥5 consecutive days^b^.

Blood and/or cerebrospinal fluid (CSF) culture positive for a pathogenic bacterial or fungal organism^c^.

^a^The terms ‘sepsis’ and ‘serious bacterial infection’ refer to clinical diagnoses of suspected or confirmed infections based on information available at the time the diagnosis is made, which may not include any laboratory test results. ‘Serious bacterial infection’ refers to an acute illness that is typically (or assumed to be) caused by bacteria such as meningitis, bacteremia/septicemia, pneumonia, urinary tract infection, osteomyelitis, septic arthritis, purulent conjunctivitis, omphalitis, pyogenic soft tissue infections, dysentery. The label ‘serious’ implies that the infection may be life-threatening or cause significant morbidity if untreated, such that conventional initial/empiric treatment in a young infant would involve parenteral antibiotics and admission to the hospital. Among infants <60 days, presumed bacterial infections are rarely considered ‘non-serious’ but, in rare cases, may include uncomplicated acute otitis media or minor/superficial skin infections.

^b^Wherever possible, the actual prescription of parenteral antibiotics was used. The need to ascertain the non-study treating physician’s intention to treat with parenteral antibiotics applied when the parenteral antibiotics prescription could not be observed accurately and/or ended early due to the parent/caregiver acting against medical advice. If an infant was removed from the hospital against medical advice, the intended (prescribed) duration of antibiotics was used to determine if this criterion was met.

^c^Severe infection episodes with culture-positive blood and/or CSF for a pathogenic bacterial or fungal organism were referred to as microbiologically-confirmed severe infection. A list of blood pathogens and contaminants was developed *a priori* by a committee with expertise in infectious diseases and microbiology and was refined after the study concluded based on the isolation of organisms that were not pre-specified (see **section S7** below).

**Section S7.** Categorization of organisms in blood as pathogens or contaminants

For both the SEPSiS Observational Cohort Study and another SEPSiS study (SEPSiS: *L. plantarum* phase II trial), a list of blood pathogens and contaminants was developed *a priori* by a committee with expertise in infectious diseases and microbiology, and *post hoc* modifications and/or expansions were made based on blood culture data generated in both studies. In brief, the list of all positive blood cultures was reviewed by a panel of two clinicians and a final determination was made regarding the categorization of each organism as a contaminant or pathogen. Each clinician was provided with a complete list of blood culture and complementary clinical data that were generated in the study of interest (i.e., the final determination of the genus and species of each organism identified, all biochemical test results that were used to identify each organism, and clinical data that were associated with the event that triggered blood collection (e.g., age of infant on day of admission to hospital, hospital admission/discharge date, signs/symptoms, diagnosis, antibiotic administration, status on discharge (alive or deceased), and duration of hospitalization).

**Categorization of organisms in blood as pathogens or contaminants**

| Categorization | Organisms |
| --- | --- |
| Pathogens | *Haemophilus influenzae*, *Streptococcus pneumoniae*, *Neisseria meningitidis*, *Escherichia coli*, *Klebsiella pneumoniae*, *Enterococcus* spp., *Enterobacter* spp., *Pseudomonas* spp., *Acinetobacter* spp., *Aeromonas* spp., *Serratia* spp., *Neisseria gonorrhea*, *Listeria monocytogenes*, Group A *streptococcus,*^a,b^ Group B *streptococcus*, Group D *streptococcus*, *Salmonella* spp., *Proteus* spp., *Citrobacter* spp., *Flavobacterium meningosepticum*, *Staphylococcus aureus*, *Morganella morganii*, *Nocardia* spp., *Moraxella* spp., *Corynebacterium jeikeium*, Shigella spp., *Campylobacter jejuni*,  *Leuconostoc* spp., *Mycobacterium* spp., *Candida* spp., *Burkholderia cepacia*^b,e^ |
| Contaminants^c^ | Coagulase-negative *Staphylococcus* spp., Other *Bacillus* spp., *Micrococcus* spp., *Corynebacterium* spp., *Propionibacterium* spp., diphtheroids, other *Neisseria* spp., viridans group streptococci, *Kocuria spp.*^d,e^ |

^a^Child Health Research Foundation (CHRF) microbiologist reported as *Streptococcus pyogenes* which is another name for *Group A streptococcus*.

^b^*Burkholderia cepacia* and *Streptococcus pyogenes* were identified using conventional culture techniques from a single blood sample that was collected from one participant enrolled in the SEPSiS: *L. plantarum* phase II trial who was clinically very ill. Isolation of two organisms from a single blood sample raised suspicion for contamination, but given the clinical severity of the infant's illness, based on review and adjudication by a panel with clinical expertise in infectious diseases, it was decided that these organisms should be categorized as pathogens in this case.

^c^Eight contaminants were identified and all were associated with cases that met criteria for severe infection using the primary definition.

^d^*Kocuria spp.* is a ubiquitous, commensal skin microbe that was identified in blood cultures from samples collected from two different infants. Based on what is known about this organism, the expert panel agreed that this organism should be categorized as a contaminant.

^e^These organisms were not in the list of pathogens and contaminants developed *a priori* in the SEPSiS observational cohort study protocol, so were categorized post-hoc.

**Section S8**. At-risk period definition

For definitions of severe infection that could be ascertained using caregiver recall and medical records (e.g., the primary definition of severe infection, culture-confirmed severe infection), the infant was still considered at-risk during temporary periods of absence from the catchment area. This is because study personnel retrospectively ascertained hospitalization and severe infection events at the next study visit using caregiver recall and medical records, even if the infant was outside the catchment area at the time of the event. For variations in case definitions of severe infection that required study personnel to ascertain all components of the case definition (e.g., checklists of clinical signs such as WHO pSBI, WHO CSI), the infant was not considered at-risk during their absence from the catchment area.

The infant-day was the smallest measured unit of person-time recorded. Each discrete at-risk period’s duration (in days) was calculated as the difference between an *exit date* and *entry date* (based on calendar dates), whereby *exit* refers to a transition from at-risk to not at-risk, and *entry* refers to the opposite transition from not at-risk to at-risk. For all infants, the initial entry date (day 0 of the at-risk period) was the date of enrolment. For each severe infection episode using the primary definition, the exit date for the at-risk period was the date on which all four of the following criteria were met: 1) SEPSiS CSI/sepsis/SBI criteria were ascertained; 2) there was administration of at least one dose of parenteral antibiotics; 3) the infant was admitted to hospital, already in hospital, or there was recommendation to admit the infant to hospital; 4) the infant’s age was less than or equal to 60 days. The day on which all four criteria were met was recorded as *day 1* of a possible severe infection event, following which subsequent consecutive days of parenteral antibiotics were counted. One day was added to person-time contributed by all infants to ensure that all infants contributed at least one day of person-time at risk (this approach was not specified in the analysis plan in the SEPSiS observational cohort study protocol, but was implemented to enable inclusion of infants who met severe infection criteria on the day of enrolment).^1^ Infants hospitalized for reasons other than severe infection (e.g., jaundice) continued to accrue person-time at risk if they were able to be visited daily in hospital by a study medical officer. The last day of a severe infection episode was the date on which the infant received (or was intended to receive) the last prescribed dose of antibiotics as part of the course of therapy for severe infection and/or the day of discharge from the hospital, whichever was later. For an infant for whom a severe infection hospitalization and antibiotic course ended early against medical advice, the last day of the severe infection episode was the non-study treating physician-prescribed duration of the antibiotic course (for the primary severe infection definition, a minimum of five consecutive days was required). For the primary analysis, the entry date for the next at-risk period was on the third severe infection-free day after the last day of a severe infection episode. Therefore, within three days of the last day of a severe infection episode, an infant was not considered to be at risk for a subsequent incident severe infection episode. For variations in severe infection case definitions (which do not include administration of antibiotics as part of the definition), infants re-entered at-risk status on or after the third severe infection-free day following hospital discharge for a prior severe infection episode.

A recurrence of a severe infection episode (as opposed to a subsequent incident severe infection episode) was defined as documentation of all four ‘day 1’ severe infection criteria more than one day after the last day on which parenteral antibiotics were administered for a prior severe infection episode (i.e., the antibiotic course was interrupted for at least one day) or the day of discharge (whichever is later), but within the 3-day period during which the infant was considered not at risk of an incident episode. Recurrences of severe infection episodes were not included in the count of severe infection episodes.

A subsequent incident severe infection episode was defined as documentation of all four ‘day 1’ severe infection criteria after the infant had re-entered at-risk status, which occurred on or after the third severe infection-free day following the last day on which parenteral antibiotics were administered (or hospital discharge, if later than the last day of parental antibiotics) for a prior severe infection episode. Subsequent incident severe infection episodes were included in the count of severe infection episodes.

**Section S9.** Sensitivity analyses

To explore the impact of informative censoring due to loss to follow-up or withdrawal from the study, a sensitivity analysis was conducted that was restricted to infants with at least 30 days of follow-up time.

We also examined the sensitivity of the incidence rate to variations in the categorization of manually entered free-text SBI diagnoses and variations in the duration of the severe infection-free period following a severe infection episode which is used to determine the re-entry into the subsequent at-risk period.

For infants who remained hospitalized following a severe infection episode, a sensitivity analysis was conducted in which the entry date for the next at-risk period was on the third severe infection-free day after the last day on which parenteral antibiotics were administered for severe infection, even if the infant remained in hospital.

A sensitivity analysis was conducted in which one day was added to person-time contributed by infants who had a severe infection episode (as opposed to adding one day to person-time contributed by all enrolled infants in the primary analysis).

Day 60 was used as the upper limit of the observation period because a routine postnatal visit was scheduled on day 60 of life. Moreover, since events occurring at non-study hospitals relied on retrospective ascertainment of hospitalization and severe infection events at the next study visit using caregiver recall and medical records, it was important to include the day-60 routine visit in the observation period to ascertain events that occurred up to day 59 of age at a non-study hospital. However, since the WHO defines the young infant age group as 0 to 59 days, a sensitivity analysis was conducted using an observation period of 0 to 59 days.

Regarding loss to follow-up, bias may be introduced by including follow-up time to the latest date of contact (scheduled or unscheduled) with study personnel because unscheduled visits to the hospital may be more likely to occur among infants who experience severe infection versus infants without severe infection. This approach may lead to an increased likelihood of including additional person-time in infants who experience severe infection versus infants without severe infection, which would increase the incidence. Using the last scheduled date of contact with study personnel as the date of loss to follow-up would mitigate this bias, but severe infection events that occurred after the last scheduled date of contact would be excluded. We conducted a sensitivity analysis using the last scheduled date of contact with study personnel.

**Table S2.** Summary of analyses that were independently verified and reviewed

| **Analysis** | **Type of review** |
| --- | --- |
| Severe infection case identification using primary definition, time at risk, and incidence estimates | Independent verification |
| Baseline characteristics of cohort, laboratory investigation results | Code review |
| Case identification using variations in severe infection definitions, time at risk, and incidence estimates | Code review |
| Sensitivity analyses | Code review |

The primary analyses, including identifying severe infection episodes, time at risk, and calculating incidence estimates, were conducted by one data analyst (CH) and underwent independent verification by a second analyst (C-YC). The second analyst was provided with instructions on how to manage the data and derive variables and statistical methods to use, but independently recoded the analysis. Discrepancies were discussed and reconciled until the same results were reached by each analyst. Code for all other analyses was written by one analyst (CH) and reviewed by a second analyst (C-YC) once the independent verification was concluded.

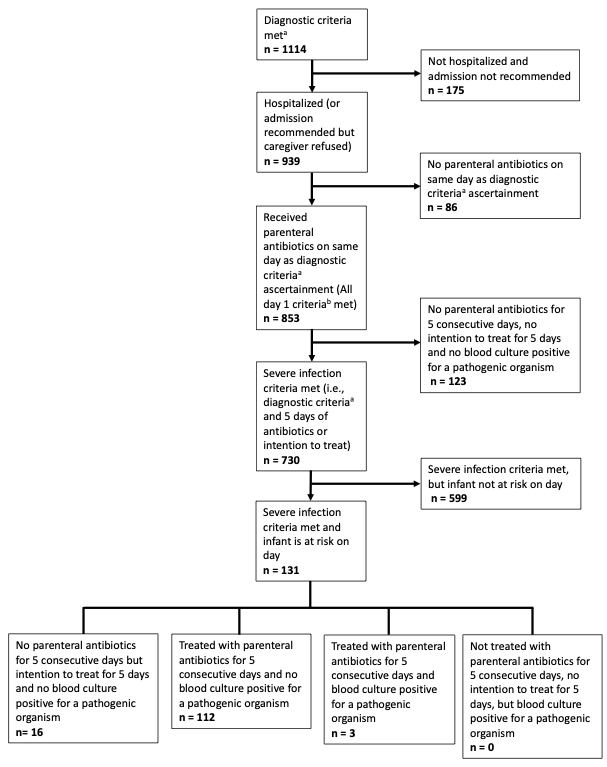

^a^Diagnostic criteria: SEPSiS Clinical Severe Infection (CSI)/Sepsis/Serious Bacterial Infection (SBI) criteria were ascertained.

^b^Day 1 criteria: 1) Diagnostic criteria were ascertained; 2) there was administration of at least one dose of parenteral antibiotics; 3) the infant was admitted to hospital, already in hospital, or there was recommendation to admit the infant to hospital; 4) the infant’s age was less than or equal to 60 days.

**Figure S2**. Flow diagram from diagnostic criteria to severe infection criteria, within the first 60 days after birth, in infant days (N_infants_=1939)

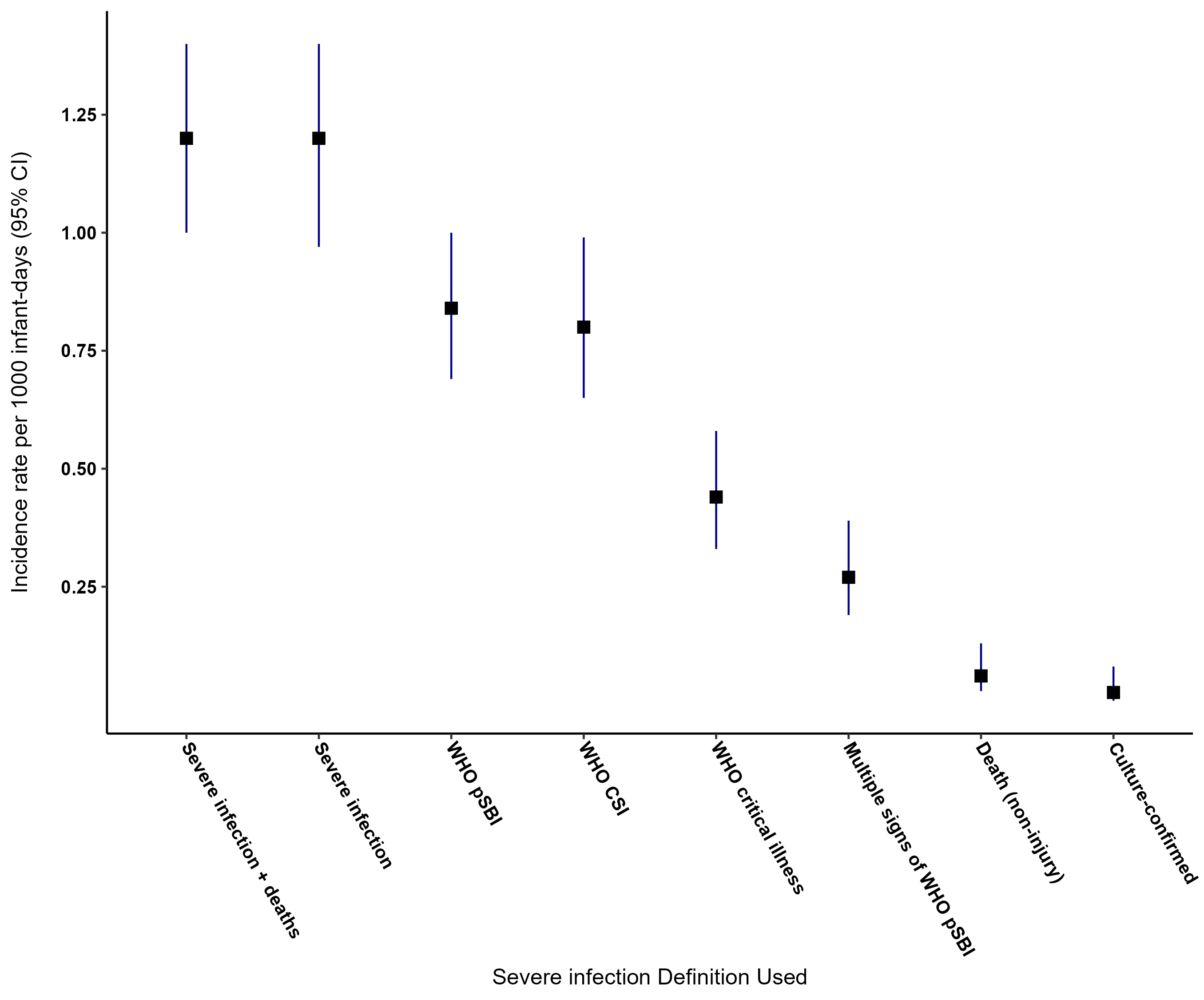

CSI: clinical severe infection; pSBI: possible serious bacterial infection; WHO: World Health Organization.

**Figure S3**. Coefficient plot comparing incidence rate estimates of severe infection and variations in severe infection case definitions

*Stratification analyses by gestational age and age at onset of event*

**Table S3.** Incidence proportions and incidence rates of severe infection by gestational age

| **Subgroup** | **Subgroup** | **Count of events** | **Number of infants with at least one event** | **Time at risk, infant-days** | **Incidence proportion, per 1000 infants at risk (95% CI)** | **Incidence rate, per 1000 infant-days (95% CI)** |
| --- | --- | --- | --- | --- | --- | --- |
| **All GA** | **Primary severe infection definition** | 131 | 128 | 113238 | 69 (58, 81) | 1.2 (0.97, 1.4) |
| **GA ≥37 weeks** | **Primary severe infection definition** | 114 | 112 | 101811 | 68 (55, 80) | 1.1 (0.93, 1.3) |
|  | **WHO possible serious bacterial infection** | 79 | 78 | 98205 | 52 (40, 64) | 0.80 (0.65, 1.0) |
|  | **WHO clinical severe infection** | 76 | 75 | 98215 | 50 (38, 61) | 0.77 (0.62, 0.97) |
| **GA <37 weeks** | **Primary severe infection definition** | 13 | 12 | 9561 | 76 (34, 117) | 1.4 (0.76, 2.4) |
|  | **WHO possible serious bacterial infection** | 11 | 10 | 9266 | 68 (26, 109) | 1.2 (0.63, 2.2) |
|  | **WHO clinical severe infection** | 10 | 10 | 9267 | 68 (26, 109) | 1.1 (0.58, 2.0) |

GA: gestational age; WHO: World Health Organization.

33 (1.7%) infants had missing gestational age data. Infants with missing gestational age data were included in the ‘All GA’ category but not the disaggregated categories.

**Table S4.** Incidence proportions and incidence rates of severe infection by age at onset of event

| **Subgroup** | **Definition** | **Count of events** | **Number of infants with at least one event** | **Time at risk, infant-days** | **Incidence proportion, per 1000 infants at risk (95% CI)** | **Incidence rate, per 1000 infant-days (95% CI)** |
| --- | --- | --- | --- | --- | --- | --- |
| **0 to 60 days of age** | **Primary severe infection definition** | 131 | 128 | 113238 | 69 (58, 81) | 1.2 (0.97, 1.4) |
| **<28 days of age** | **Primary severe infection definition** | 78 | 78 | 51048 | 43 (34, 52) | 1.5 (1.2, 1.9) |
|  | **WHO possible serious bacterial infection** | 49 | 48 | 50458 | 29 (20, 37) | 0.97 (0.73, 1.3) |
|  | **WHO clinical severe infection** | 45 | 45 | 50492 | 27 (19, 35) | 0.89 (0.67, 1.2) |
| **28 to 60 days of age** | **Primary severe infection definition** | 53 | 52 | 62304 | 27 (20, 35) | 0.85 (0.65, 1.1) |
|  | **WHO possible serious bacterial infection** | 43 | 42 | 57804 | 22 (15, 29) | 0.74 (0.55, 1.0) |
|  | **WHO clinical severe infection** | 43 | 42 | 59702 | 24 (17, 31) | 0.72 (0.53, 0.98) |

**Table S5**. Incidence proportions and incidence rates of severe infection by sex

| **Subgroup** | **Definition** | **Count of events** | **Number of infants with at least one event** | **Time at risk, infant-days** | **Incidence proportion, per 1000 infants at risk (95% CI)** | **Incidence rate, per 1000 infant-days (95% CI)** |
| --- | --- | --- | --- | --- | --- | --- |
| **All** | **Primary severe infection definition** | 131 | 128 | 113238 | 69 (58, 81) | 1.2 (0.97, 1.4) |
| **Female** | **Primary severe infection definition** | 68 | 66 | 59279 | 68 (52, 84) | 1.1 (0.9, 1.5) |
|  | **WHO possible serious bacterial infection** | 45 | 45 | 57124 | 53 (37, 69) | 0.79 (0.59, 1.1) |
|  | **WHO clinical severe infection** | 44 | 44 | 57135 | 52 (36, 68) | 0.77 (0.58, 1.0) |
| **Male** | **Primary severe infection definition** | 63 | 62 | 53959 | 70 (53, 87) | 1.2 (0.91, 1.5) |
|  | **WHO possible serious bacterial infection** | 47 | 45 | 52192 | 54 (38, 69) | 0.90 (0.67, 1.2) |
|  | **WHO clinical severe infection** | 44 | 43 | 52192 | 51 (36, 66) | 0.84 (0.63, 1.1) |

**Table S6**. Sensitivity analyses for primary severe infection case definition incidence proportion and rate

|  | **Count of events** | **Number of infants with at least one event** | **Time at risk, infant-days** | **Incidence proportion, per 1000 infants at risk (95% CI)** | **Incidence rate, per 1000 infant-days at risk (95% CI)** |
| --- | --- | --- | --- | --- | --- |
| **Primary severe infection definition** | 131 | 128 | 113238 | 69 (58, 81) | 1.2 (0.97, 1.4) |
| **Return to at-risk the day after last day of a severe infection episode** | 132 | 128 | 113463 | 69 (58, 81) | 1.2 (0.97, 1.4) |
| **Return to at-risk 10 days after last day of a severe infection episode** | 130 | 128 | 112445 | 69 (58, 81) | 1.2 (0.97, 1.4) |
| **‘Likely a serious bacterial infection (SBI)’ diagnoses only** | 129 | 126 | 113257 | 68 (57, 80) | 1.1 (0.96, 1.4) |
| **Add 1 day at-risk to severe infection cases only rather than to all infants** | 131 | 128 | 111427 | 69 (58, 81) | 1.2 (0.99, 1.4) |
| **Only infants with at least 30 days of follow-up time** | 129 | 126 | 112959 | 68 (57, 80) | 1.1 (0.96, 1.4) |
| **Last day of severe infection as last day of antibiotics only (not last day of antibiotics or last day of discharge, whichever is later)** | 131 | 128 | 113299 | 69 (58, 81) | 1.2 (0.97, 1.4) |
| **Using observation period of 0-59 days** | 128 | 125 | 111370 | 68 (56, 79) | 1.1 (0.97, 1.4) |
| **Using last scheduled date of contact with study personnel (rather than using latest scheduled or unscheduled date of contact)** | 131 | 128 | 113202 | 69 (58, 81) | 1.2 (0.98, 1.4) |

**Table S7**. Laboratory investigation results

| **Laboratory investigation** | **Threshold for clinical concern^a^** | **Median (25^th^, 75^th^)** | **Number of severe infection episodes for which test was done (%)^b^, N=131** | **Number of severe infection episodes with test result meeting threshold for clinical concern (%)** | **Additional information** |
| --- | --- | --- | --- | --- | --- |
| White Blood Cell Count (10^9^/L) | >15 | 11 (8.7, 13) | 87 (66) | 7 (8.0)^c^ | N/A |
| Neutrophil Count (10^9^/L) | >6.75 | 3.5 (2.0, 4.9) | 87 (66) | 14 (16)^d^ | N/A |
| Platelet Count (10^9^/L) | >500 | 387 (289, 489) | 87 (66) | 19 (22)^e^ | N/A |
| **High-Sensitivity C-Reactive Protein (mg/L)** | >5 | 1.5 (0.45, 4.8) | 83 (63) | 20 (24) | N/A |
| **Procalcitonin (ng/mL)** | ≥0.5 | 0.11 (0.070, 0.44) | 79 (60) | 17 (22) | N/A |
| **Glucose (mmol/L)** | <3.3 | 4.8 (3.9, 5.6) | 77 (59) | 13 (17) | N/A |
| **Creatinine (micromol/L)** | >73 | 27 (20, 36) | 81 (62) | 6 (7.4) | N/A |
| **Alanine Aminotransferase (U/L)** | >30 | 17 (14, 23) | 83 (63) | 4 (4.8) | N/A |
| **Total Bilirubin (micromol/L)^f^** | Day of life 0: >200 Day of life 1: >265 Day of life 2: >300 Day of life 3: >340 >Day of life 3: >360 | 68 (15, 169) | 84 (64) | 1 (1.2) | N/A |
| **Urine Dipstick** | Positive for leukocyte esterase and/or nitrites | N/A | 85 (65) | 15 (18) | Positive for leukocyte esterase (15 episodes)  Positive for nitrites (0 episodes)  Positive for leukocyte esterase and nitrites (0 episodes) |
| **RT-PCR Nasal swabs** | Positive for a pathogenic organism | N/A | 111 (85) | 35 (32) | RSV (30 episodes)  Influenza (2 episodes)  *Ureaplasma* (1 episode)  SARS-CoV2 (1 episode)  RSV and *Ureaplasma* (1 episode) |
| **Skin swabs^g^** |  | N/A | 4 (3.1) | 4 (100) | *Staphylococcus aureus* (4 episodes)^h^ |
| **Urine culture** |  | N/A | 87 (66) | 8 (9.2) | *E. coli* (1 episode)  *Enterococcus* spp. (6 episodes)  *Enterobacter cloacae* (1 episode) |
| **Blood culture** |  | N/A | 105 (80) | 3 (2.9) | *S. aureus* (2 episodes) *Citrobacter freundii* (1 episode) |

^a^Thresholds based on combination of clinical judgment of two physicians on SEPSiS team and Hospital for Sick Children Guide to Lab Services reference range values.

^b^Proportions of severe infection episodes for which tests were done are below 100% for tests that should have been collected for all severe infection episodes. This may have been due to low blood volumes collected and the need to prioritize testing for some investigations over others.

^c^Number of severe infection episodes with white blood cell count <5.0 (10^9^/L) (%) was 7 (8.0).

^d^Number of severe infection episodes with neutrophil count <0.5 (10^9^/L) (%) was 3 (3.4).

^e^Number of severe infection episodes with platelet count <150 (10^9^/L) (%) was 3 (3.4).

^f^Total bilirubin is not a test that raises clinical concern for severe infection but was included in in the study design. Bilirubin values were reported back to study medical officers who then referred infants for care if values were outside of normal ranges for age.

^g^Skin swabs were done only if there was clinical concern for skin infection, and swabs were taken of the infected area.

^h^Three of the staphylococcus aureus-positive skin swabs were penicillin-resistant and one was penicillin-sensitive. Data were not available on sensitivity to methicillin.

**Table S8.** Positive cultures captured by the primary *severe infection* definition and the WHO *possible serious bacterial infection* (pSBI) definition

|  | Blood culture | Urine culture | Skin swab cultures from infected area |
| --- | --- | --- | --- |
| Number of samples positive for a pathogenic organism in SEPSiS cohort | 3 | 8 | 4 |
| Severe infection | 3 | 8 | 4 |
| WHO possible serious bacterial infection | 2 | 6 | 0 |

**Table S9**. Characterization of severe infection episodes by RSV/influenza positivity, biomarkers, urine dipstick results and urine and blood culture positivity

| **Clinical characterization of episode** | **Laboratory criteria** | **Number of severe infection episodes for which test was done (%), N=131** | **Number of severe infection episodes with test result meeting threshold for clinical concern (%)** |
| --- | --- | --- | --- |
| Probable viral and/or bacterial infection | [RSV/influenza positive]^a^  OR  [Evidence of a systemic inflammatory response^b^]  OR  [Bacterial culture^c^ positive or urine dipstick positive^d^] | 67 (51) | 47 (70) |
| Probable viral illness without bacterial infection | [RSV/Influenza positive^a^ and bacterial culture^c^ negative]  OR  [No evidence of a systemic inflammatory response^b^]  AND  [Bacterial culture^c^ and urine dipstick negative^e^] | 67 (51) | 45 (67) |
| Possible or confirmed bacterial infection (without evidence of viral etiology) | [RSV/Influenza negative]^a^  AND  [Evidence of systemic inflammatory response^b^]  AND/OR  [Bacterial culture^c^ positive or urine dipstick positive^d^] | 67 (51) | 24 (36) |

^a^Of the panel of nasal swab pathogens, only respiratory syncytial virus (RSV) and influenza A and B were included in the characterization of severe infection episodes because these viruses were considered to be likely causes of the presenting illness.

^b^Evidence of a systemic inflammatory response = white blood cell count, neutrophil count, C-reactive protein, or procalcitonin meet threshold for clinical concern. Threshold for clinical concern includes being beyond either lower or upper limits of the reference range. If any of white blood cell count, neutrophil count, C-reactive protein, or procalcitonin were done, the test of the composite of ‘white blood cell count, neutrophils, C-reactive protein, or procalcitonin’ was considered to have been done.

^c^Bacterial culture = urine or blood culture

^d^Urine dipstick positive = positive for leukocyte esterase and/or nitrites

^e^Urine dipstick negative = negative for leukocyte esterase and negative for nitrites

**Table S10**. Sample size calculations for a theoretical randomized controlled trial of a severe infection prevention intervention in young infants

By rate:^2^

$$Equation 1: n infants required =\left[ \frac{\left( 1.96+0.84 \right)^{2}\left( \mu1+ \mu0 \right)}{\left( \mu1- \mu0 \right)^{2}} \right]/60$$

*µ*0: Incidence rate of severe infection or alternative severe infection definition

*µ*1: *µ*0 × Incidence rate ratio

Number of infants *per group* of trial needed to detect a difference at α=0.05, β=0.2 assuming all infants were observed from 0-60 days.

| Incidence rate ratio ($\mu1/\mu0)$ | Severe infection  ($\mu0= 1.2$) | WHO possible serious bacterial infection  $(\mu0= 0.84)$ | Non-injury death  $(\mu0= 0.061)$ | Culture-confirmed infection  ($\mu0=0.026)$ |
| --- | --- | --- | --- | --- |
| 0.90 | 20,689 | 29,556 | 406,995 | 954,872 |
| 0.80 | 4,900 | 7,000 | 96,393 | 226,154 |

By proportion:^2^

$$Equation 2: n infants = \frac{\left( 1.96+0.84 \right)^{2}* (\pi0*(1-\pi0))+(\pi1*(1-\pi1))}{{(\pi0 - \pi1)}^{2}}$$

*π*0: Incidence proportion of severe infection or alternative severe infection definition

*π*1: π0 × Relative risk

Number of infants *per group* of trial needed to detect a difference at α=0.05, β=0.2.

| Relative risk ($\pi1/\pi0)$ | Severe infection  $\pi0=0.066$ | WHO possible serious bacterial infection  $\pi0=0.046$ | Non-injury death  $\pi0= 0.0036$ | Culture confirmed infection  $\pi0=0.015$ |
| --- | --- | --- | --- | --- |
| 0.90 | 21,151 | 30,964 | 412,359 | 991,648 |
| 0.80 | 5,024 | 7,348 | 97,679 | 234,879 |
